# Analysis and Prediction of COVID-19 Patients’ False Negative Results for SARS-CoV-2 Detection with Pharyngeal Swab Specimen: A Retrospective Study

**DOI:** 10.1101/2020.03.26.20043042

**Authors:** Hui Xu, Li Yan, Chun (Martin) Qiu, Bo Jiao, Yanyan Chen, Xi Tan, Zhuo Chen, Ling Ai, Yaru Xiao, Ailin Luo, Shusheng Li

## Abstract

**Background:** False negative results of SARS-CoV-2 nucleic acid detection pose threats to COVID-19 patients and medical workers alike.

**Objective:** To develop multivariate models to determine clinical characteristics that contribute to false negative results of SARS-CoV-2 nucleic acid detection, and use them to predict false negative results as well as time windows for testing positive.

**Design:** Retrospective Cohort Study (Ethics number of Tongji Hospital: No. IRBID: TJ-20200320)

**Setting:** A database of outpatients in Tongji Hospital (University Hospital) from 15 January 2020 to 19 February 2020.

**Patients:** 1,324 outpatients with COVID-19

**Measurements:** Clinical information on CT imaging reports, blood routine tests, and clinic symptoms were collected. A multivariate logistic regression was used to explain and predict false negative testing results of SARS-CoV-2 detection. A multivariate accelerated failure model was used to analyze and predict delayed time windows for testing positive.

**Results:** Of the 1,324 outpatients who diagnosed of COVID-19, 633 patients tested positive in their first SARS-CoV-2 nucleic acid test (47.8%), with a mean age of 51 years (SD=14.9); the rest, which had a mean age of 47 years (SD=15.4), tested negative in the first test. “Ground glass opacity” in a CT imaging report was associated with a lower chance of false negatives (aOR, 0.56), and reduced the length of time window for testing positive by 26%. “Consolidation” was associated with a higher chance of false negatives (aOR, 1.57), and extended the length of time window for testing positive by 44%. In blood routine tests, basophils (aOR, 1.28) and eosinophils (aOR, 1.29) were associated with a higher chance of false negatives, and were found to extend the time window for testing positive by 23% and 41%, respectively. Age and gender also affected the significantly.

**Limitation:** Data were generated in a large single-center study.

**Conclusion:** Testing outcome and positive window of SARS-CoV-2 detection for COVID-19 patients were associated with CT imaging results, blood routine tests, and clinical symptoms. Taking into account relevant information in CT imaging reports, blood routine tests, and clinical symptoms helped reduce a false negative testing outcome. The predictive AFT model, what we believe to be one of the first statistical models for predicting time window of SARS-CoV-2 detection, could help clinicians improve the accuracy and efficiency of the diagnosis, and hence, optimizes the timing of nucleic acid detection and alleviates the shortage of nucleic acid detection kits around the world.

**Primary Funding Source:** None.

## Introduction

COVID-19, the disease caused by SARS-CoV-2 virus that surfaced in Wuhan, China in early December 2019, is now plaguing the world. Due to its high intra-human transmission nature, there were more than one hundred and fifty thousand confirmed cases around the world by 15 March, 2020. In view of the rapid surge of infected patients, World Health Organization (WHO) has declared the viral disease a pandemic on 11 March, 2020.

To contain COVID-19, a prompt and accurate diagnostic test is necessary. While new tests have been proposed, the SARS-CoV-2 nucleic acid test is currently the standard diagnostic criterion used by most countries. However, contradictory to clinical symptoms and chest CT scanning, many patients have shown false negative testing results during their initial clinic visits. As a result, hospital admission and treatment have been delayed for many of them. These patients have been stranded in outpatient clinics or isolation zones, increasing the risk of exposure of other non-COVID-19 patients and medical workers. Their confirmed diagnoses of COVID-19 have since raised concerns for the validity and reliability of the test. To better contain COVID-19, it is essential to understand the factors that influence the test’s false-negative incidence rate.

Several factors contribute to a relatively high false-negative incidence in the nucleic acid test: (1) the sensitivity of the detection kits; (2) inappropriate clinical sampling from patients; (3) the original viral load. The first factor is a concern for the manufacturers while the second can be remedied through staff training. The last factor, however, links to the progression of COVID-19, which is patient-specific. In other words, the viral load has demonstrated individual heterogeneity, and has gone beyond doctors’ subjective evaluation. Hence, it is essential to conduct the test when viral load is high. Consequently, the right timing for the test will avoid repeated sampling, reduce exposure risk for outpatients and healthcare providers, and improve the efficiency of medical efforts to contain the epidemic.

This study retrospectively analyzed the relationships between clinical characteristics and the nucleic acid test results of COVID-19 patients; it developed statistical models to predict nucleic acid test results for patients diagnosed with COVID-19: how likely they are to test positive, and when they are likely to test positive. Based on a patient’s clinical characteristics, the study proposed a model to predict a time window that would help doctors identify the right time for testing. The findings shed light on the development for more accurate and efficient clinical diagnosis procedures for COVID-19, and may alleviate the shortage of nucleic acid detection kits around the world.

## Methods

### Study population and data collection

This single-centered, retrospective study was approved by the Human Assurance Committee (HAC) of Tongji Hospital (No. IRBID: TJ-20200320), (affiliated with Tongji Medical College, Huazhong University of Science and Technology, Wuhan, China). We collected the electronic records of 11,368 outpatients in Tongji Fever Clinic through standardized data collection tables in the electronic medical records. A total of 3,588 patients, from the 11,368 outpatients, who underwent the SARS-CoV-2 nucleic acid test were screened from 15 January 2020 to 19 February 2020. Among them, 2,264 patients were excluded due to the unavailability of their CT imaging reports and/or blood routine tests. Finally, 1,324 patients diagnosed with COVID-19 were enrolled for this study (633 patients tested positive in their first nucleic acid test, and the rest 691 patients tested negative initially, and subsequently tested positive later on). Their epidemiological information, including age and gender, CT imaging reports, clinical symptoms, and blood routine test results, nucleic acid test results were processed for analysis.

### Nucleic acid assay

Laboratory confirmation of SARS-CoV-2 was conducted as follows: pharyngeal swab specimens from the upper respiratory tract were collected from outpatients. The swab was placed into a collection tube with virus preservation solution. Total RNA was extracted using the respiratory sample RNA isolation kit approved by the Food and Drug Administration of China. Two target genes, including the open reading frame 1ab (ORF1ab) and the nucleocapsid protein (N) of SARS-CoV-2, were simultaneously amplified by Real-time reverse transcriptase– polymerase chain reaction (rRT-PCR) as previously described (1).

### Variables of interest

The variables for which we intend to seek explanations and make predictions are: 1) outcome of the first nucleic acid test for diagnosed COVID-19 patients; and 2) time duration between getting sick and testing positive.

Based on previous research that studied the clinic characteristics and diagnosis of COVID-19 (1-3), we assembled a list of characteristics from three information sources: CT imaging reports, clinic visit records, and blood routine tests to explain and predict the above two variables of interest. Six key important initial visit symptoms were extracted from clinic visit records and were coded as dummy variables (0 if such a symptom was not reported; and 1 if reported). Ten blood routine test items were collected, four of which were identified as diagnostic indicators via pilot analysis. They were lymphocytes, basophils, eosinophils, and neutrophils (all in counts). Through the text mining approach, a number of key phrases from CT imaging reports (in Chinese) were generated. Based on the findings in recent studies (4, 5), six of which were chosen to be included in the analyses. They were translated as “ground glass opacity (hereafter GGO)”, “patchy shadows”, “subsolid”, “consolidation”, “bilateral pulmonary”, and “unilateral pulmonary”. They were also coded as dummy variables (0 if such a characteristic was not detected; and 1 if detected). The text mining task was conducted via an R package called “JiebaR” for text mining in Chinese.

### Initial probing

We followed the norm in the time-to-event analysis (aka survival analysis), where the event is defined as a positive test result in the nucleic acid test. The time window is defined as the duration between the time of a patient getting sick and the time of the event (first positive test result) taking place. Since the time of testing positive was not the exact time the patient reached the threshold of enough viral load for testing positive, the time window was indeed interval censored. Some patients’ test results remained negative during the data collection. For those patients, their time windows were treated as right censored following the standard treatment in the survival analysis.

### Statistical analysis

Statistical tests were performed using R statistical software (version 3.3.6). Two different multivariate analyses were employed to retrospectively decompose the effects on the nucleic acid test results of patients diagnosed with COVID-19.

One analysis used logistic regression to study how clinical characteristics were associated with the test results of diagnosed patients. The dependent variable for the logistic regression was the outcome of a patient’s first nucleic acid test (negative vs. positive). The analysis was conducted via an R command “glm”.

The other analysis used the accelerated failure time (AFT) models. AFT models investigated the time-to-event window by linking the time window of testing positive to clinical characteristics. The dependent variable was the logarithm of the time difference between getting sick and testing positive. Specifically, the AFT model is specified as

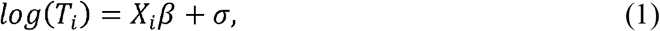

where *X*_*i*_ include factors of CT imaging reports, clinic visit records, and blood tests introduced above. The distribution of the error term *σ* is assumed to be exponential. An R package “survival” was used, with the “survreg” function to estimate AFT models.

All reported *P* values were two-sided; and all reported results bear a statistical significance with a *P* value less than 0.05.

### Prediction

The predictions of nucleic acid test outcomes for diagnosed patients were conducted in two different aspects: the on-the-spot outcome (positive vs. negative) for a patient’s first nucleic acid test; and a time window for testing positive. In both aspects, we divided the data into the testing and validation samples by a ratio of 80%-20%. We trained the model on the testing sample and generated predicted probabilities of false negative test result on the validation sample via the logistic regression. We then compared the predicted probabilities with actual test results and plotted ROC curves.

Alternatively, the length of the time window for testing positive was predicted via the AFT model. We formulated the AFT model analysis with all three types of clinical characteristics (CT imaging, blood test results, and clinical symptoms) for prediction. Once we estimated the coefficients on the training sample, we obtained a set of estimates 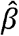. For any patient in the validation sample (with index *k*), we use equation (2) below to calculate the predicted probabilities of testing negative for a given time *t*, given by

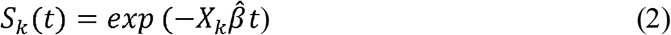

Since each patient in the validation sample was given a predictive curve that linked the probabilities of testing negative and the timespan since getting sick, we then calculated the cut-off timespan where the probability of testing negative, *S*(*t*), started to become smaller than 50%:

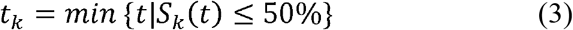

We took into consideration the confidence intervals of *t*_*k*_ to construct the time window for testing positive. For those patients eventually testing positive in the data, the predicted time window is given as (*t*_*k*_ − *cσ*_*k*_, +∞), where *σ*_*k*_ is the standard error, and *c* is a constant equal to 1.96 for the 95% confidence interval. For patients whose positive test results were censored (the exact time for testing positive is unavailable in the data), the time window for testing negative is given as (0, *t*_*k*_ + *cσ*_*k*_). We then calculated the percentage of the patients whose timespan for the test correctly matched their predicted time window. The prediction proposed in this study offered better interpretation and tractability than the concordance index.

## Results

### Baseline clinical characteristics

A total of 1,324 patients who were diagnosed with COVID-19 were eligible for this study. Of the 1,324 patients, 1,270 were diagnosed as having a light condition (95.9%), and the rest were diagnosed as either severe (3.8%) or critical (0.3%). 633 patients tested positive in their first nucleic acid test (47.8%) (**Table 1**).

**Table 1.** Baseline and characteristics of 1324 patients.

| **Diagnosed COVID-19** |  | **First nucleic acid test outcome** |  |  |
| --- | --- | --- | --- | --- |
| **Patients (N=1324)** |  |  |  |  |
| **Basic characteristics** | **Positive**<br>**(N=633)** | **Negative**<br>**(N=691)** | **Total**<br>**(N=1324)** | **P value<sup>†</sup>** |
| **Age, mean (SD), y** | 51 (14.9) | 47 (15.4) | 49 (15.3) | <0.001<sup>‡</sup> |
| Young ($\leq 35$) y, n(%) | 105 (16.6) | 177 (25.6) | 282 (21.3) | <0.001 |
| Middle age ($>35\text{--}\leq 55$) y, n(%) | 254 (40.1) | 288 (41.7) | 542 (40.9) | |
| Younger old ($>55\text{--}\leq 74$) y, n(%) | 247 (39.0) | 208 (30.1) | 455 (34.4) | |
| Old ($>74$) y, n(%) | 27 (4.3) | 18 (2.6) | 45 (3.4) | |
| **Gender, n(%)** |  |  |  |  |
| Female | 325 (51.3) | 348 (50.4) | 673 (50.8) | 0.721 |
| Male | 308 (48.7) | 343 (49.6) | 651 (49.2) |  |
| **Classification of disease, n(%)** |  |  |  |  |
| Light | 604 (95.4) | 666 (96.4) | 1 270 (95.9) | 0.455<sup>§</sup> |
| Severe | 26 (4.1) | 24 (3.5) | 50 (3.8) |  |
| Critical | 3 (0.5) | 1 (0.1) | 4 (0.3) |  |
| **CT image characteristics, n(%)** |  |  |  |  |
| Ground-glass opacity | 383 (60.5) | 288 (41.7) | 671 (50.7) | <0.001 |
| Patchy shadows | 412 (65.1) | 361 (52.2) | 773 (58.4) | <0.001 |
| Subsolid | 7 (1.1) | 9 (1.3) | 16 (1.2) | 0.744 |
| Consolidation | 71 (11.2) | 96 (13.9) | 167 (12.6) | 0.143 |
| Bilateral pulmonary | 533 (84.2) | 498 (72.1) | 1 031 (77.9) | <0.001 |
| Unilateral pulmonary | 284 (44.9) | 404 (58.5) | 688 (52.0) | <0.001 |
| **Initial symptoms, n(%)** |  |  |  |  |
| Fever | 507 (80.1) | 470 (68.0) | 977 (73.8) | <0.001 |
| Cough | 201 (31.8) | 215 (31.1) | 416 (31.4) | 0.802 |
| Fatigue | 84 (13.3) | 85 (12.3) | 169 (12.8) | 0.598 |
| Chest distress | 71 (11.2) | 98 (14.2) | 169 (12.8) | 0.106 |
| Diarrhea | 15 (2.4) | 13 (1.9) | 28 (2.1) | 0.537 |
| Loss of appetite | 32 (5.1) | 23 (3.3) | 55 (4.2) | 0.116 |
| **Blood routine test** |  |  |  |  |
| Lymphocytes ($\times 10^9$/L; NR: 1.1-3.2) | | | | |
| Median (IQR) | 1.21 (0.93-1.59) | 1.45 (1.07-1.93) | 1.33 (1.00-1.76) | <0.001<sup>††</sup> |
| Increased, n% | 4 (0.6) | 20 (2.9) | 24 (1.8) | <0.001 |
| Decreased, n% | 247 (39.0) | 180 (26.1) | 427 (32.3) |  |
| Basophils ($\times 10^9$/L; NR: 0-0.1) | | | | |
| Median (IQR) | 0.01 (0.00-0.01) | 0.01 (0.01-0.02) | 0.01 (0.005-0.02) | < 0.001<sup>††</sup> |
| Increased, n% | 0 (0.0) | 5 (0.7) | 5 (0.4) | 0.063<sup>§</sup> |
| Eosinophils ($\times 10^9$/L; NR: 0.02-0.52) | | | | |
| Median (IQR) | 0.01 (0.00-0.03) | 0.03 (0.005-0.08) | 0.02 (0.00-0.06) | < 0.001<sup>††</sup> |
| Increased, n% | 1 (0.2) | 6 (0.9) | 7 (0.5) | < 0.001<sup>§</sup> |
| Decreased, n% | 398 (62.9) | 263 (38.1) | 661 (49.9) |  |
| Neutrophils ($\times 10^9$/L; NR: 1.8-6.3) | | | | |
| Median (IQR) | 3.50 (2.59-4.64) | 3.77 (2.70-5.27) | 3.63 (2.63-4.91) | 0.001<sup>††</sup> |
| Increased, n% | 59 (9.3) | 102 (14.8) | 161 (12.2) | 0.009 |
| Decreased, n% | 42 (6.6) | 47 (6.8) | 89 (6.7) |  |

COVID-19 = Coronavirus Disease 2019; SD = Standard deviation; NR = Normal range; IQR = interquartile range.
<sup>†</sup>P values comparing COVID-19 positive group and negative group were from $\chi^2$ test.
<sup>‡</sup>P value was calculated by T test.
<sup>§</sup>P values were calculated by Fisher exact test.
<sup>††</sup>P values were calculated by Mann-Whitney test.

In terms of age distribution, the average age for the diagnosed patients with a positive result in the first nucleic acid test (aka FNAC-P patients) was 51 years (SD=14.9). The diagnosed patients who tested negative in the first nucleic acid test (aka FNAC-N patients) was 47 years (SD=15.4). FNAC-P patients consisted of 308 males (48.7%) and 325 females (51.3%). In comparison, FNAC-N patients consisted of 343 males (49.6%) and 348 females (50.4%) (**Table1**).

Remarkably, it is noteworthy that the typical CT images derived from FNAC-P patients were characterized by GGO and patchy shadows, which occurred at 60.5% or 65.1% of all FNAC-P patients, respectively. Only 41.7% or 52.2% of FNAC-N patients displayed the above manifestations, respectively. In contrast, the FNAC-P patients were less likely to have consolidation in their CT images than FNAC-N patients (11.2% vs. 13.9%) (**Table 1**).

Some blood routine tests also showed significant differences. The average number of lymphocytes in FNAC-P patients was 1.21×10^9^/L (IQR 0.93-1.59), while it was 1.45×10^9^ /L (IQR 1.07-1.93) for the FNAC-N patients (**Table 1**). In contrast, FNAC-P patients displayed lower numbers of eosinophils counts as compared to that of FNAC-N patients (0.01×10^9^/L (IQR 0.00-0.03) vs. 0.03×10^9^/L (IQR 0.005-0.08)). Similarly, a lower number of basophils was characterized in FNAC-P patients as compared to that of FNAC-N patients (0.01×10^9^/L (IQR 0.00-0.01) vs.0.01×10^9^/L (IQR 0.01-0.02)) (**Table 1**).

### Kaplan–Meier time-to-event curves

To echo the results in Table 1, we plotted several Kaplan–Meier time-to-event curves to illustrate the probabilities of remaining negative against the time window (in days) (**Figure 1**). The higher the position of a KM curve, the more likely were the patients to test positive given the same timespan since getting sick. In other words, they had a shorter time window for testing positive. It was noticeable that the detection of the phrase “GGO” in CT imaging reports reduced the time window for testing positive (**Figure 1 A**). In comparison, the detection of “consolidation” extended the time window for testing positive, leading to a higher chance of false negative if the test was taken in earlier time (**Figure 1 B**). It was clear that higher levels of basophils and eosinophils delayed the time for testing positive, resulting in a higher likelihood for a false negative (**Figure 1 C, D**). Among clinic symptoms, fever was the only one to be associated with the time window by shortening its duration (**Figure 1 E**), whereas chest distress was the only symptom to be associated with the time window by extending it (**Figure 1 F**). Finally, in it was shown that elder male patients were likely to experience a shorter time window for testing positive (**Figure 1 G, H**).

**Figure 1.**
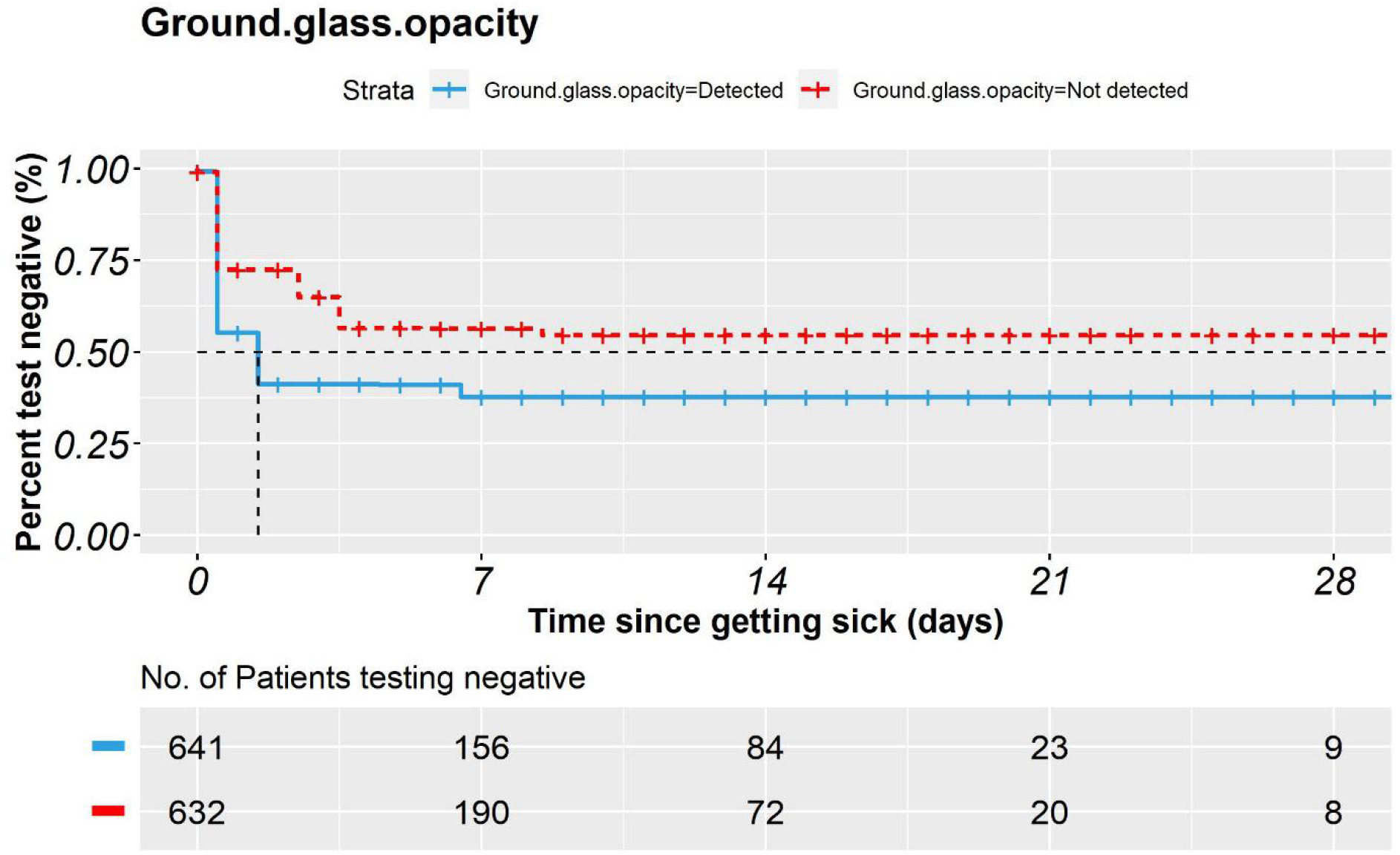
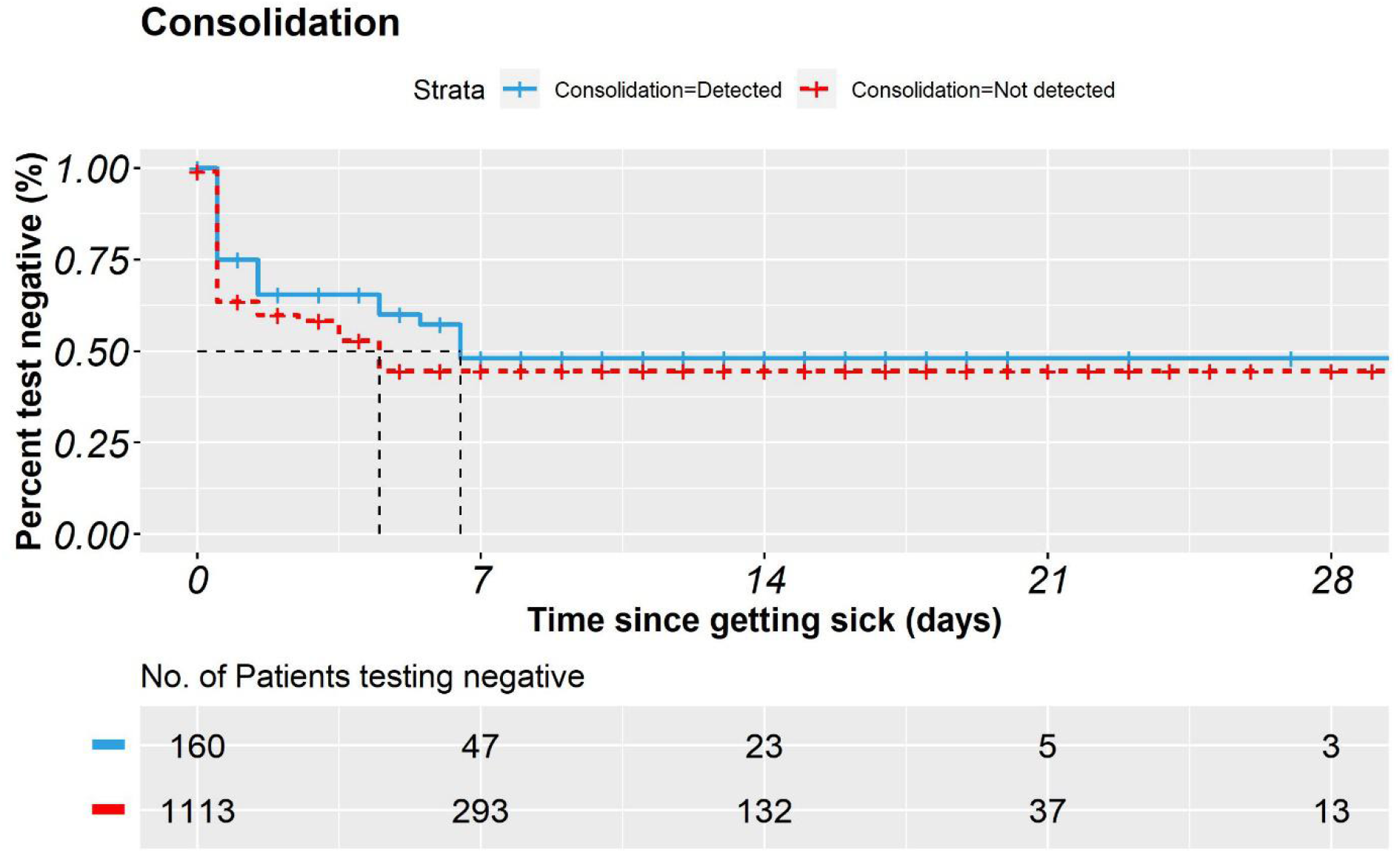
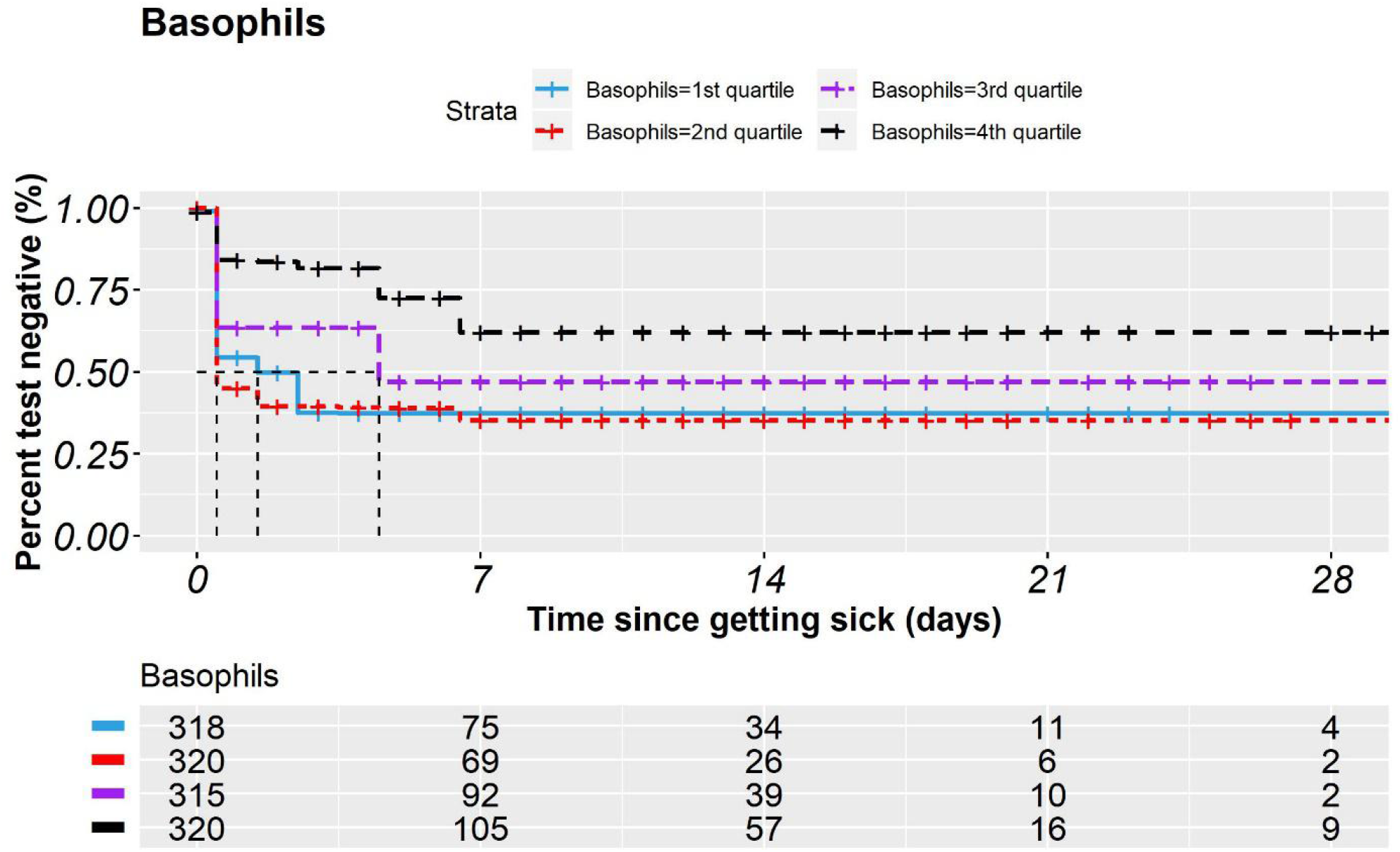
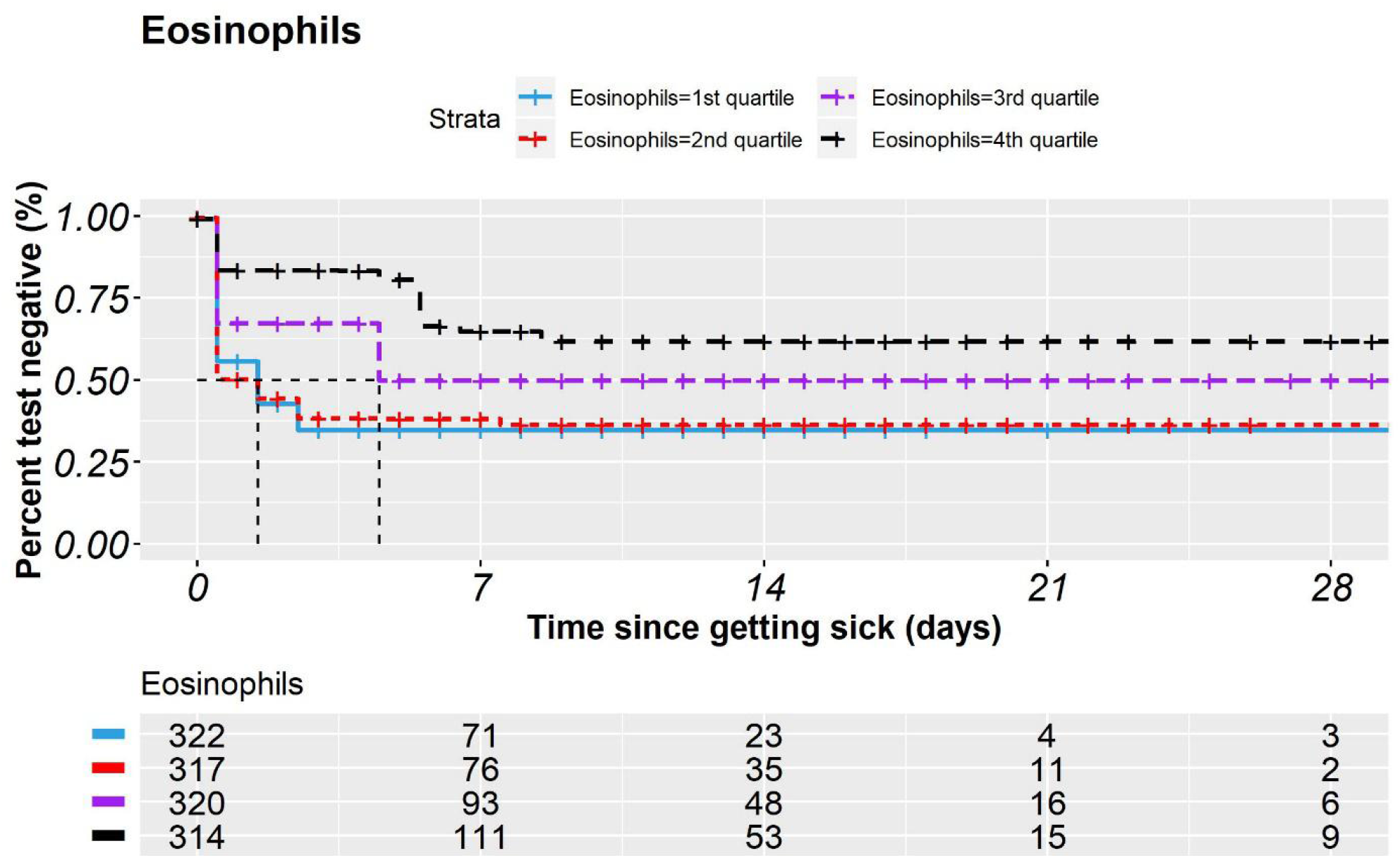
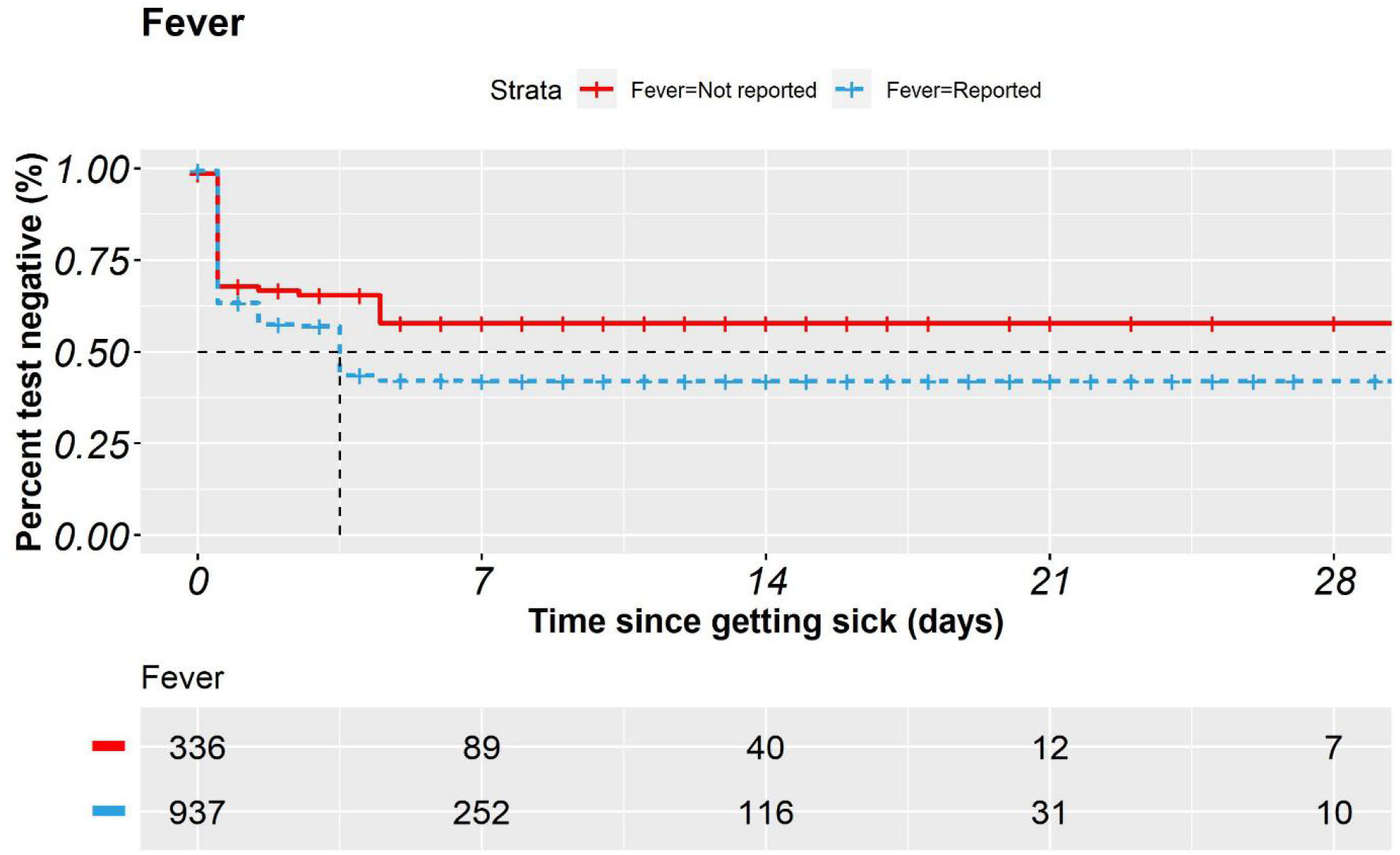
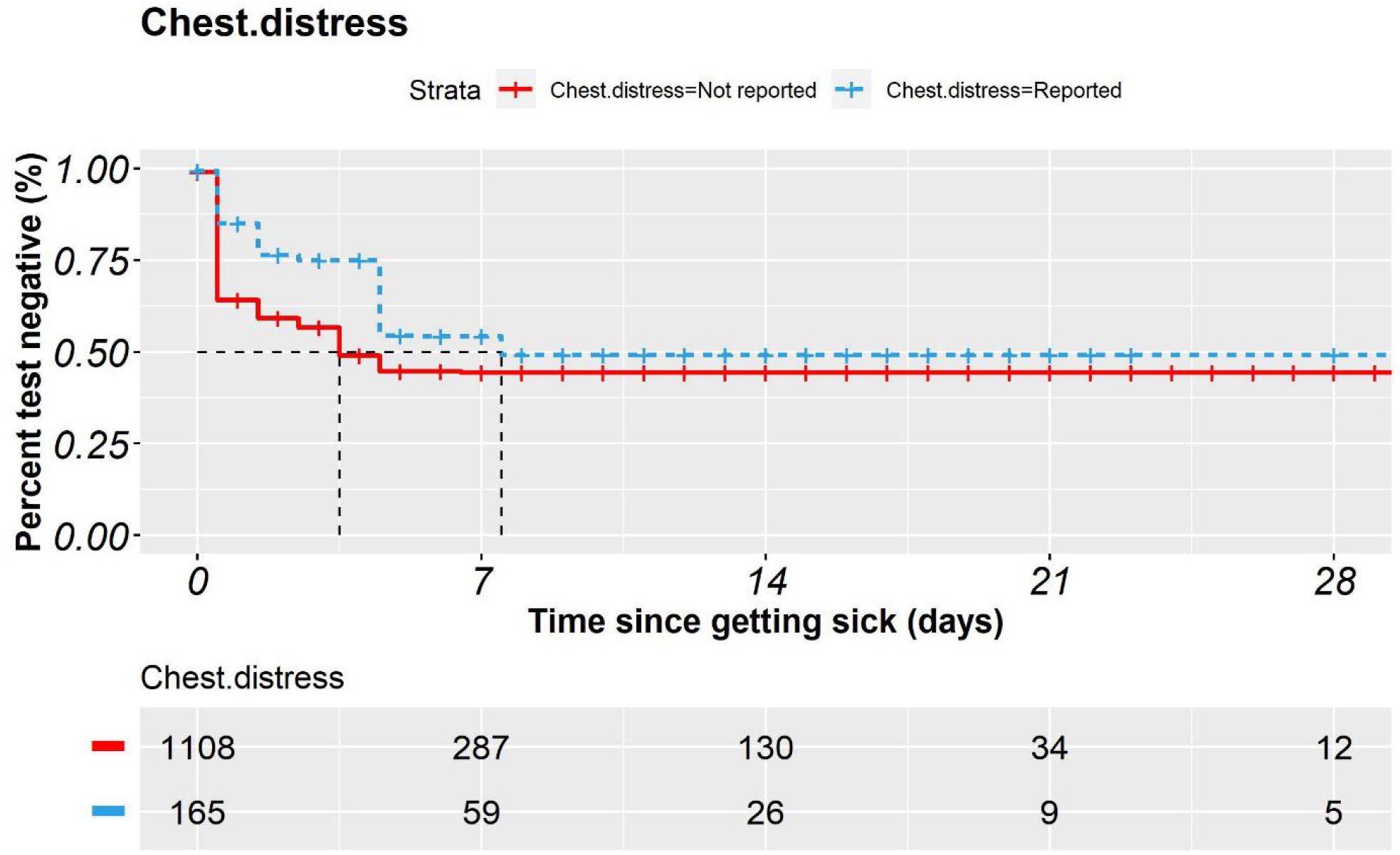
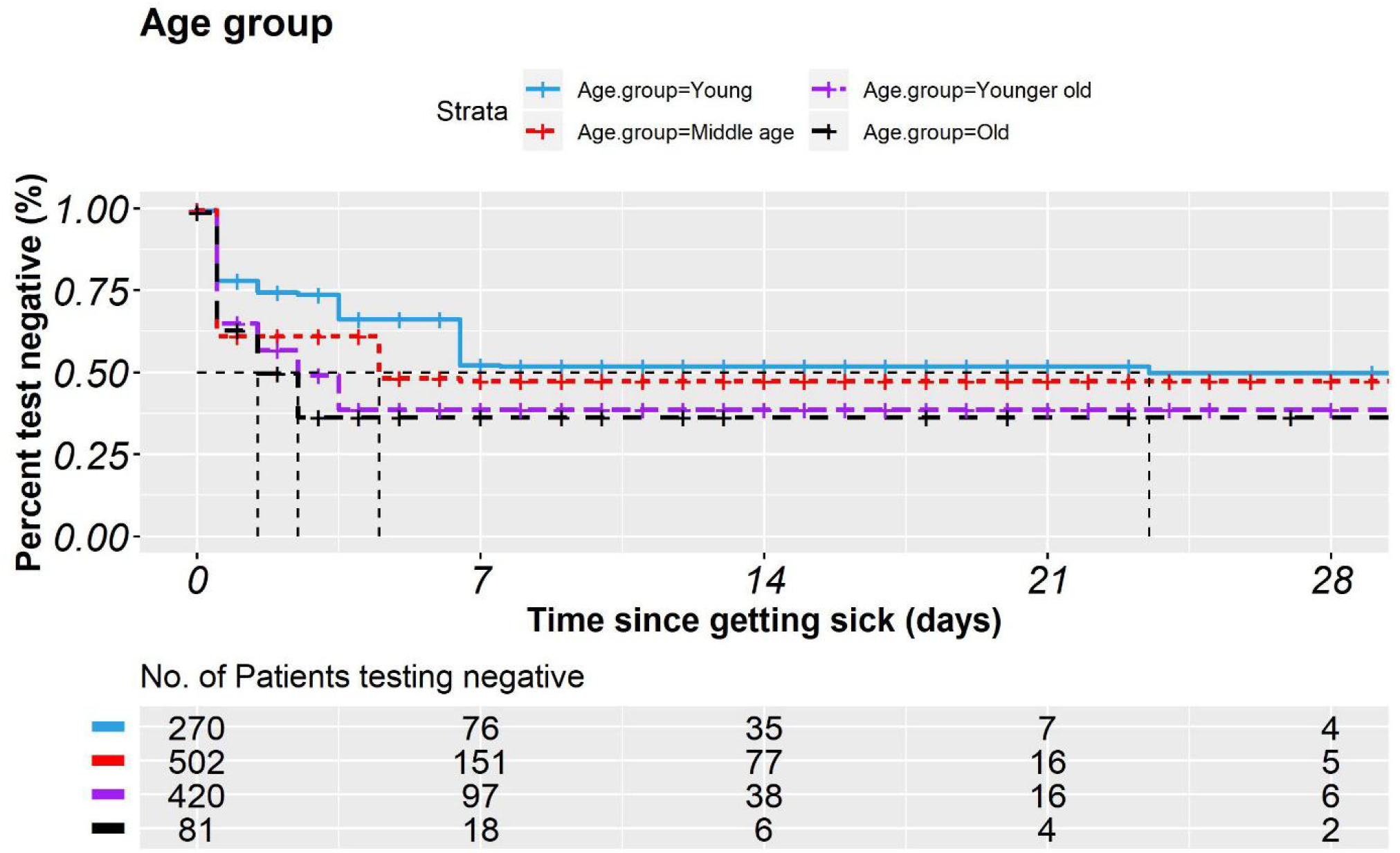
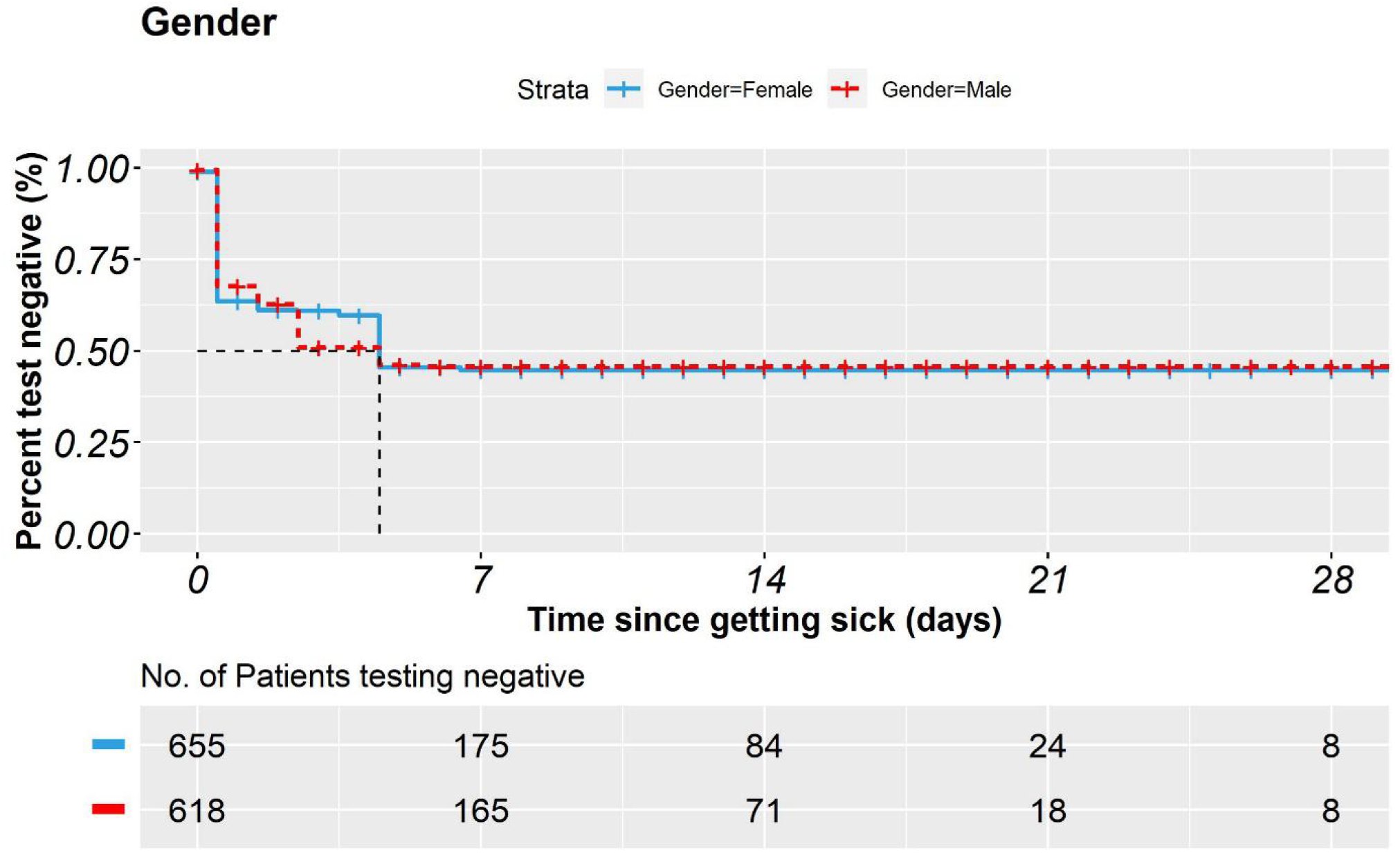
Kaplan–Meier time-to-event curves. **A**. K-M plot for ground glass opacity (GGO) detected vs. not detected in CT imaging reports. The lower detected curve showed a shorter time window for testing positive. **B**. K-M plot for consolidation detected vs. not detected in CT imaging reports. The higher detected curve showed a longer time window for testing positive. **C**. K-M plot for basophils in blood routine tests. Lab results were divided into four quartiles by the values, with the 1st quartile including 25% of patients of the lowest test results, and so on. Higher level of basophils delayed the time window for testing positive. **D**. K-M plot for eosinophils in blood routine tests. Lab results were divided into four quartiles by the values, with the 1st quartile including 25% of patients of the lowest test results, and so on. Higher level of eosinophils delayed the time window for testing positive. **E**. K-M plot for fever symptom reported in patients’ visits. Patients with fever had a shorter time window for testing positive. **F**. K-M plot for chest distress symptom reported in patients’ visits. Patients with chest distress had a longer time window for testing positive. **G**. K-M plot for four age groups difference. “Young” was defined as age ≤ 35. “Middle age” was for age in the range of (35, 55]. “Younger old” was for age in (55, 74], and “Old” was for age greater than 74. Older patients were related to a shorter time window for testing positive. **H**. K-M plot for gender difference. Male patients on average had a shorter time window for testing positive.

### Logistic regression results

The logistic regression provided factors of interest on the test outcome after the adjustment for age, gender, and timespan between getting sick and the test (**Table 2**). We found one characteristic of CT imaging reports, “GGO”, to be associated with a lower chance of false negative (adjusted odds ratio aOR, 0.56; 95% CI, 0.44-0.71; *P* <0.001). Nevertheless, two characteristics of CT imaging reports were associated with a higher chance of false negative: “consolidation” (aOR, 1.57; 95% CI, 1.08-2.27; *P* = 0.02) and “unilateral Pulmonary” (aOR, 1.46; 95% CI, 1.11-1.9; *P* = 0.01). Fever was also found to be associated with a lower chance of false negative (aOR, 0.7; 95% CI, 0.51-0.96; *P* = 0.02). Out of the blood test items, two were found to be associated with a higher chance of false negative: Basophils and (aOR, 1.28; 95% CI, 1.14-1.44; *P* < 0.001) Eosinophils (aOR, 1.29; 95% CI, 1.03-1.62; *P* = 0.03). These findings remained consistent when we used different model specifications.

**Table 2.**
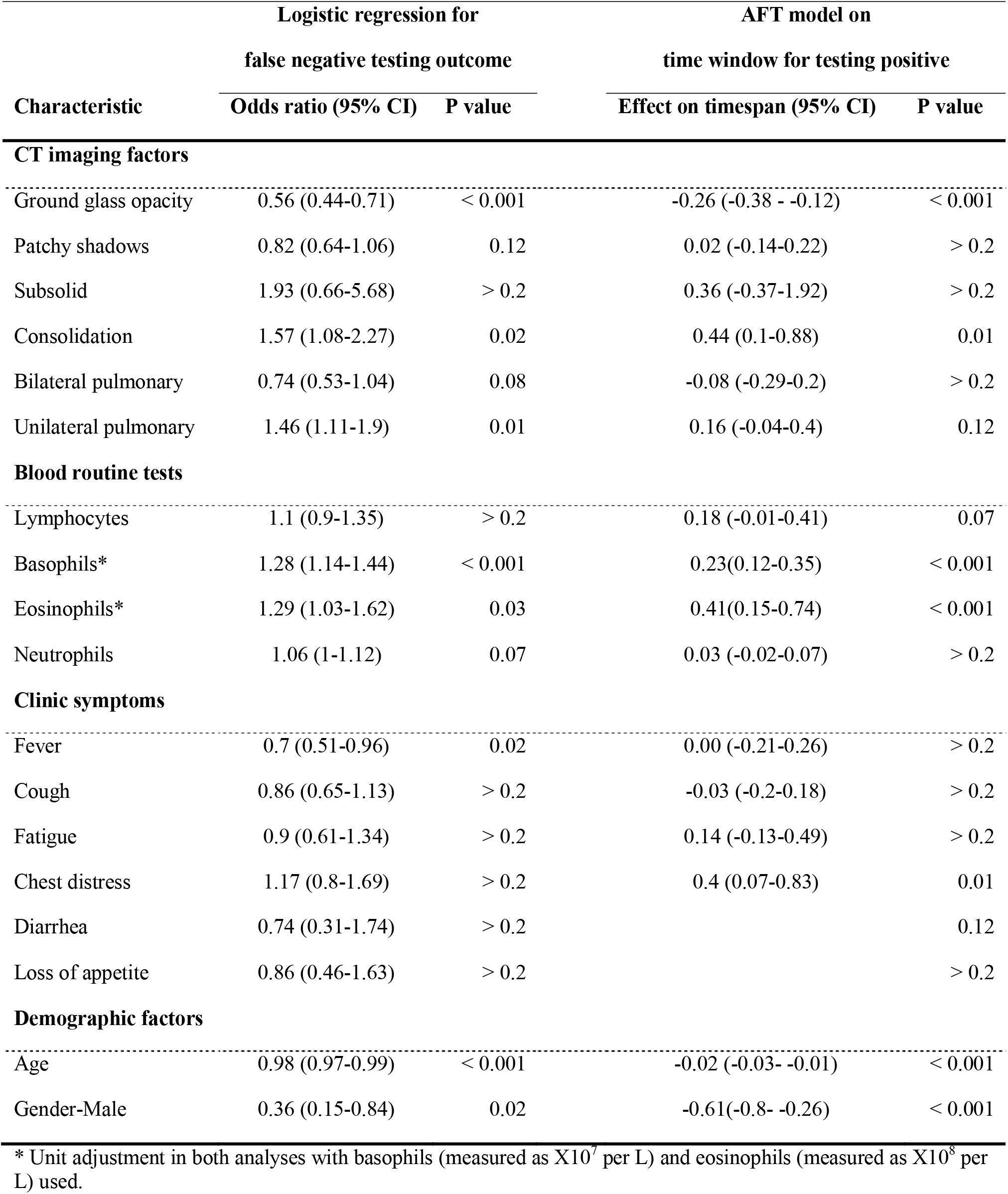
Estimation results for logistic regression and accelerated failure time (AFT) model.

| Characteristic | Logistic regression for<br>false negative testing outcome |  | AFT model on<br>time window for testing positive |  |
| --- | --- | --- | --- | --- |
|  | Odds ratio (95% CI) | P value | Effect on timespan (95% CI) | P value |
| **CT imaging factors** |  |  |  |  |
| Ground glass opacity | 0.56 (0.44-0.71) | < 0.001 | -0.26 (-0.38 - -0.12) | < 0.001 |
| Patchy shadows | 0.82 (0.64-1.06) | 0.12 | 0.02 (-0.14-0.22) | > 0.2 |
| Subsolid | 1.93 (0.66-5.68) | > 0.2 | 0.36 (-0.37-1.92) | > 0.2 |
| Consolidation | 1.57 (1.08-2.27) | 0.02 | 0.44 (0.1-0.88) | 0.01 |
| Bilateral pulmonary | 0.74 (0.53-1.04) | 0.08 | -0.08 (-0.29-0.2) | > 0.2 |
| Unilateral pulmonary | 1.46 (1.11-1.9) | 0.01 | 0.16 (-0.04-0.4) | 0.12 |
| **Blood routine tests** |  |  |  |  |
| Lymphocytes | 1.1 (0.9-1.35) | > 0.2 | 0.18 (-0.01-0.41) | 0.07 |
| Basophils* | 1.28 (1.14-1.44) | < 0.001 | 0.23(0.12-0.35) | < 0.001 |
| Eosinophils* | 1.29 (1.03-1.62) | 0.03 | 0.41(0.15-0.74) | < 0.001 |
| Neutrophils | 1.06 (1-1.12) | 0.07 | 0.03 (-0.02-0.07) | > 0.2 |
| **Clinic symptoms** |  |  |  |  |
| Fever | 0.7 (0.51-0.96) | 0.02 | 0.00 (-0.21-0.26) | > 0.2 |
| Cough | 0.86 (0.65-1.13) | > 0.2 | -0.03 (-0.2-0.18) | > 0.2 |
| Fatigue | 0.9 (0.61-1.34) | > 0.2 | 0.14 (-0.13-0.49) | > 0.2 |
| Chest distress | 1.17 (0.8-1.69) | > 0.2 | 0.4 (0.07-0.83) | 0.01 |
| Diarrhea | 0.74 (0.31-1.74) | > 0.2 |  | 0.12 |
| Loss of appetite | 0.86 (0.46-1.63) | > 0.2 |  | > 0.2 |
| **Demographic factors** |  |  |  |  |
| Age | 0.98 (0.97-0.99) | < 0.001 | -0.02 (-0.03- -0.01) | < 0.001 |
| Gender-Male | 0.36 (0.15-0.84) | 0.02 | -0.61(-0.8- -0.26) | < 0.001 |

\* Unit adjustment in both analyses with basophils (measured as $\times 10^7$ per L) and eosinophils (measured as $\times 10^8$ per L) used.

In order to implement the out-of-sample prediction, 1,324 patients were randomly divided into training (1,059 cases) and validation samples (265 cases). Three different ROC curves were plotted for three different model specifications of the logistic regression (**Figure 2**). AUC values ranged from 0.69 to 0.77.

**Figure 2.**
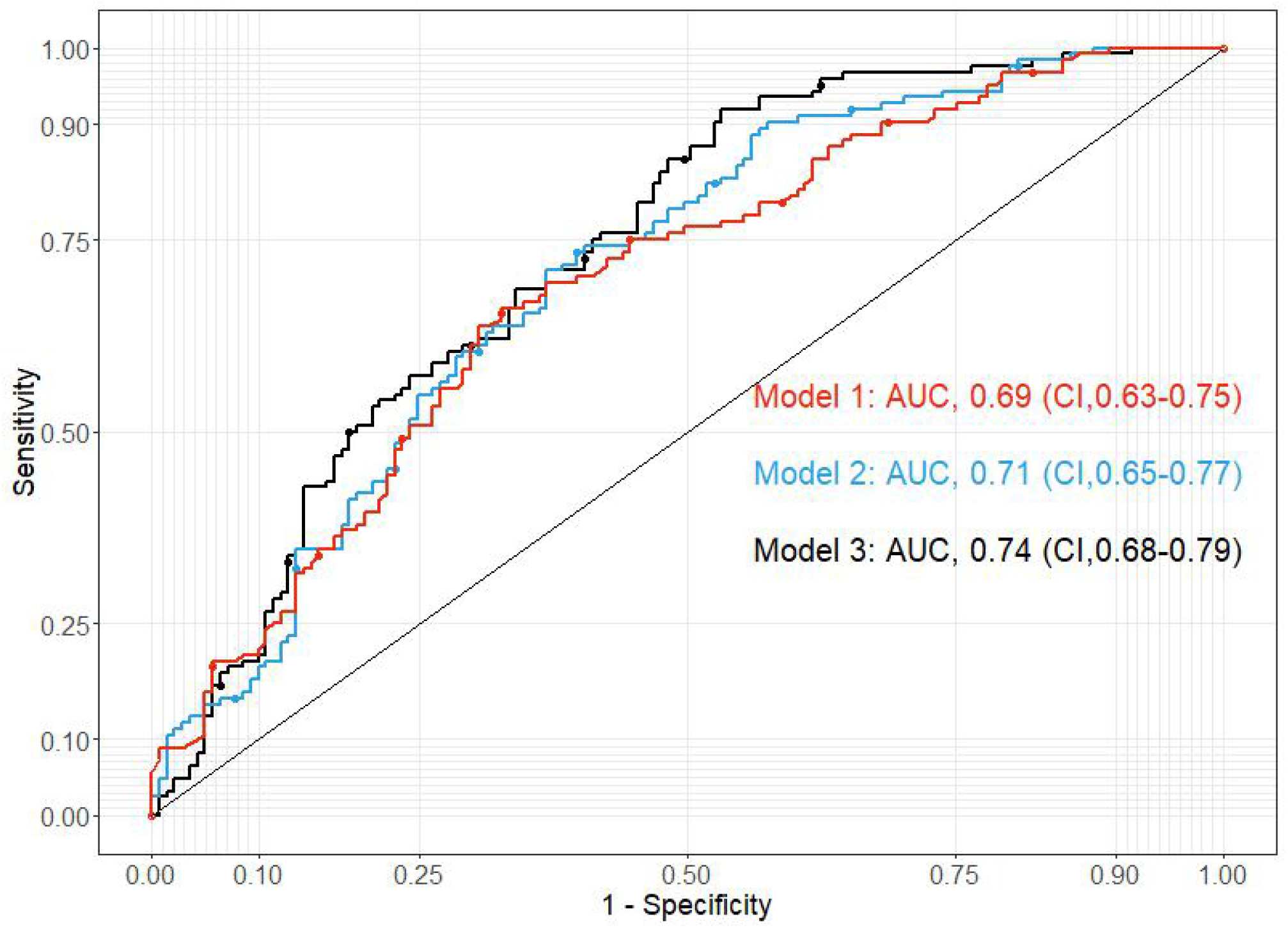
Receiver operating characteristic curves of three different model specifications. Receiver operating characteristic curves of three different model specifications are plotted. Model 1 included six CT imaging characteristics, controlling for age and gender. Model 2 included all the factors in model 1, plus six clinical symptoms and time gap between getting sick and CT/tests. Model 3 included all the factors in model 2, plus four blood routine test items.

### Accelerated failure time (AFT) model results

The AFT model was used to calibrate the effects of factors of interests on the length of the time window of testing positive. The coefficient estimates from the AFT analysis provided the impact of a particular characteristic on the length of time window (in percentage) under a multivariate environment, controlling for other factors (**Table 2**). “GGO” was associated with a shorter window for testing positive: “GGO” (effect, -0.26, 95% CI, -0.38--0.12; *P* < 0.001), suggesting the detection of this characteristic will on average reduce the length of the time window of testing positive by 26%. In comparison, the detection of “consolidation” (effect, 0.44; 95% CI, 0.1-0.88; *P* = 0.01) will on average extend the length of the time window by 44%. The results are consistent with the logistic regression. We also found that chest distress was associated with a longer window as well (effect, 0.4; 95% CI, 0.07-0.83; *P* = 0.01). Finally, two blood test items, basophils (effect, 0.23; 95% CI, 0.12-0.35; *P* < 0.001) and eosinophils (effect, 0.41; 95% CI, 0.15-0.74; *P* < 0.001) were also linked to a longer time window for testing positive, implying that higher values of test results supressed the positive testing outcome. These results were also consistent with the logistic regression.

### Time window prediction and validation

The AFT model was used to predict each patient’s time window for testing positive. The training and validation were conducted on the same testing and validation samples. To illustrate the prediction process, we used two patients as examples (**Figure 3**).

**Figure 3.**
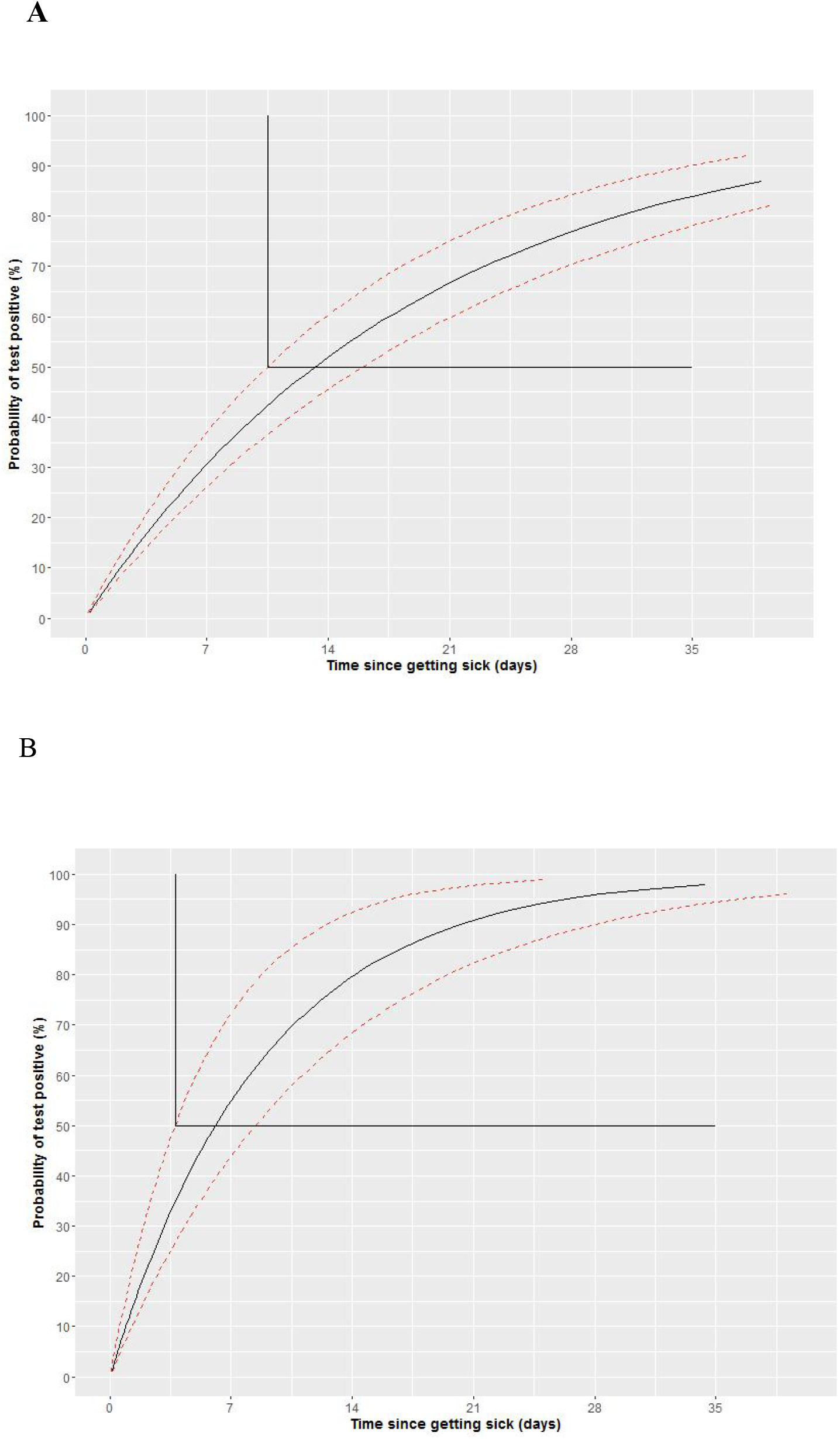
Predicting positive testing outcome windows for diagnosed patients. For a 37-year-old male patient with consolidation detected but not ground glass opacity in his CT imaging report, fever reported from his clinic visit record, and the blood routine test results (lymphocytes: 1.68×10^9^/L, basophils: 0.01×10^9^/L, eosinophils: 0.04×10^9^/L and neutrophils: 2.56×10^9^/L), it was predicted that the time window for testing positive started at 10.49 days since getting sick. The patient took the test on day 21, and he had a chance greater than 50% to test positive. For a 66-year-old female patient with both ground glass opacity and consolidation detected in her CT imaging report, fever reported from her clinic visit record, and the blood routine tests results (lymphocytes: 1.07×10^9^/L, basophils: 0.015×10^9^/L, eosinophils: 0.05×10^9^/L and neutrophils: 5.65×10^9^/L), it was predicted that the time window for testing positive started at 3.77 days since getting sick. The patient took the test on day 11, so she had a chance greater than 50% to test positive.

Patient A was a 37-year-old male. He had consolidation but not GGO detected in his CT imaging report, reported fever, and his blood routine test showed the following results: lymphocytes: 1.68×10^9^/L, basophils: 0.01×10^9^/L, eosinophils: 0.04×10^9^/L and neutrophils: 2.56 ×10^9^/L. It was predicted that his time window for testing positive started at 10.49 days since getting sick. The patient took the test on day 21, and he had a chance greater than 50% to test positive (75.1%), which matched his actual test result (**Figure 3 A**).

Patient B was a 66-year-old female with both GGO and consolidation detected in her CT imaging report. She also reported fever, and her blood routine test results were: lymphocytes: 1.07×10^9^/L, basophils: 0.015×10^9^/L, eosinophils: 0.05×10^9^/L and neutrophils: 5.65×10^9^/L. It was predicted that her time window for testing positive started at 3.77 days since getting sick. Thus, having GGO detected significantly shortened the time window for testing positive, as predicted in the AFT model (**Table 2**). The patient took the test on day 11, so she had a chance greater than 50% to test positive (82.3%), as what happened to her (**Figure 3 B**).

We obtained the prediction outcome for all the patients in validation samples. Out of 265 patients, 133 patients tested positive in the data. The predicted time window correctly matched 87 patients’ actual testing time. For 132 patients who remained testing negative falsely in the sample, the predicted time window correctly matched 114 patients’ actual testing time. The overall accuracy is 75.8% (201/265).

## Discussion

COVID-19 is an acute infectious disease caused by SARS-CoV-2 infection. The standard diagnosis relies on the nucleic acid test (rRT-PCR) of the virus. Recent studies reported that higher viral loads in the swab were detected soon after symptom onset (6, 7). In the fever clinic of Tongji Hospital, we found that for many patients, whose CT images showed consolidations or diffuse infections, had more false negative nucleic acid tests. Maybe the viral load in the upper respiratory tract determines the time window for testing positive during this different course of COVID-19. The level and duration of infectious virus replication are important factors in assessing the risk of transmission and guiding decisions regarding isolation of patients. For diagnosed COVID-19 patients, a failure of early confirmation through the nucleic acid test could be disastrous. This study aimed to develop multivariate models to explain false negative test results, and further to predict a time window for testing positive. In other words, we also provided direct answers to the variety in the time windows of COVID-19 patients (8).

A CT scan is critical in helping doctors diagnose COVID-19 patients as a clear destruction of the pulmonary parenchyma is a typical result (9, 10). In both the logistic regression and the AFT model, several CT imaging characteristics were found to play a key role in affecting test results. Specifically, both GGO and consolidation were identified as key characteristics, but they worked in an opposite manner. As mentioned above, some patients presented characteristic radiographic features of COVID-19 from the first scan, while the nucleic acid test result was falsely negative (11). And then these patients were confirmed by positive repeat swab testing during the isolated observation or treatment. From our study, we found that there was a higher incidence rate of GGO in FNAC-P patients than in FNAC-N patients. GGO was the main radiological demonstration in the early stage of COVID-19 (12), and the time windows depicted in Figures 3 A and 3B highlight the importance of GGO in bringing in a shorter time window for testing positive. Our finding is also consistent with a recent study (13) that found GGO was linked to inflammatory lesions (Grayish-white lesion under naked eye). Combined with the multivariate analyses in our study, it suggests both GGO and consolidation should both serve as a heads-up for doctors to interpret test results.

This study also identified age as a key indicator for testing positive. While age was found to play an important role in susceptibility and prognosis of COVID-19 patients (1, 2, 14), this study identified that older patients were more prone to testing positive, and had a shorter window for doing so. This result was consistent with Liu (15). It further revealed that the age effect did not fade as time elapsed.

While previous research showed that both genders are susceptible to SARS-CoV-2 virus infection(1, 3), this study found that gender difference between FNAC-P and FNAC-N patients was statistically significant. Although gender was not significant in the univariate context, in the multivariate context, the effect of gender was significant and robust, as both the logistic regression and the AFT model provided strong evidence to support it.

This study also prompts us to think about whether the current quarantine standards for discharging patients are correct. According to the guidelines for the diagnosis and treatment of COVID-19 in China, when a patient tested negative twice in the nucleic acid test (an interval of 1-2 days), the patient could go home. However, one recent study reported that the detectable SARS-CoV-2 RNA persisted for a median of 20 days in survivors and that it was sustained until death in nonsurvivors (8). And some patients were later found to test positive after discharge. Some researchers believe that recovered patients may still carry the virus (16); and that the viral load of some patients may not accumulate to a high level enough for testing positive. While others believe that sampling, patient stages, reagent sources, laboratory operations, and other factors could affect test outcome. Our models help detect a delayed time window of testing positive with the analysis of some clinical characteristics and explained the variety in the time windows of COVID-19 patients. As a result, medical staff should sample at an appropriate time, taking clinical characteristics into consideration, so as to reduce false negatives.

Several limitations should be considered when interpreting these findings. Firstly, this is a single center, and retrospective study. The results need more multiple, prospective research to verify it. Secondly, with the exception of the time window, there are other reasons related to false negative results. For example, it has been reported that the sensitivity of SARS-CoV-2 nucleic acid detection of sputum specimens is higher than that of pharyngeal swabs (7, 17), which may be related to the main invasion of SARS-CoV-2 on lower respiratory tract cells. However, a dry cough is the main manifestation of COVID-19 patients, imposing difficulty on obtaining sputum specimens. With an unsatisfactory liquefaction of sputum specimen, false negative nucleic acid results increase. In addition, the collection of sputum specimen from the lower respiratory tract is easy to cause spatter, which increases the risk of infection for the operator, so it is not recommended to be used in an outpatient clinic.

In conclusion, we present what we believe to be one of the first statistical models for predicting nucleic acid test results for patients diagnosed with COVID-19. The predictive model of time window for testing positive could help clinicians identify patients at a higher risk and improve the rate of accurate diagnosis of COVID-19. The model can be extended to predict false negative of other tests for SARS-CoV-2. More external validation studies are now required to demonstrate predictions in diverse patient populations. In our follow-up studies, further refinement of this model could be achieved by including novel clinical predictors.

## Data Availability

The data that support the findings of this study are available from the corresponding author upon reasonable request.

From Department of Anesthesiology, Tongji Hospital of Tongji Medical College, Huazhong University of Science and Technology, Wuhan, Hubei, China (H.X, B.J, X.T, L.A, AL.L); Department of Emergency, Tongji Hospital of Tongji Medical College, Huazhong University of Science and Technology, Wuhan, Hubei, China(L.Y, YR.X, SS.L); Lazaridis School, Wilfrid Laurier University, Waterloo, Ontario(M.Q); Department of Information Management, Tongji Hospital of Tongji Medical College, Huazhong University of Science and Technology, Wuhan, Hubei, China(Y. Ch); Outpatient department office, Tongji Hospital of Tongji Medical College, Huazhong University of Science and Technology, Wuhan, Hubei, China(Z. Ch)

## Acknowledgments

We thank all the patients and their families involved in this study, as well as all the doctors, nurses and volunteers who working together fighting against COVID-19 in Hubei.

## Reproducible Research Statement

Study protocol and statistical code: Available from Dr. Xu (e-mail). Data set: Not available.

